# Reduced Neural Convergence and Reorganized Paranoia-Related Representations During Ambiguous Social Narrative Processing in Schizophrenia

**DOI:** 10.64898/2026.09.16.26362988

**Authors:** Lin Zhang, Bihua Xu, Anfu Deng, Jiayu Feng, Yanlong Hou, Shaolei Guo, Feng Zhou, Yulin Wang, Jiaxi Liao, Debo Dong

## Abstract

Schizophrenia may involve a fundamental disruption in the ability to construct a shared understanding of social reality, yet conventional neuroimaging studies have largely examined social cognition using simplified and highly structured paradigms that may not capture how individuals interpret complex and ambiguous interpersonal events in everyday life. Using an ecologically valid naturalistic fMRI paradigm, we examined shared and paranoia-related neural representations during ambiguous social narrative processing in 38 patients with schizophrenia and 46 healthy controls. Intersubject correlation was used to quantify neural convergence across individuals, whereas intersubject representational similarity analysis was used to examine how individual differences in paranoid ideation were reflected in neural representations. Compared with healthy controls, patients with schizophrenia showed reduced neural convergence within distributed systems supporting social inference, contextual integration, and salience attribution, including temporoparietal, temporal, insular, and inferior frontal regions. Furthermore, paranoid ideation was associated with altered neural representational structure. In healthy controls, higher paranoia was linked to reduced neural similarity across social-cognitive and affective regions, suggesting reduced convergence toward shared interpretations of ambiguous social information. In schizophrenia, paranoia-related representations showed sparse within-group associations but elevated patient–control differences across a distributed social-affective circuit, indicating a reorganization rather than a simple amplification of paranoia-related processing. Together, these findings demonstrate that schizophrenia involves both impaired alignment of neural responses during shared social meaning construction and organization of paranoia-related representations, providing an ecologically grounded framework for understanding abnormal social inference in psychosis.

## Introduction

Understanding social events is a social process that is impacted by shared meanings and values (Maitlis 2005; Kuhlmann et al. 2023). Individuals frequently rely on a set of shared contextual conventions to evaluate the behaviors of others, infer their intentions, and integrate a few social cues into a coherent narrative (Hasson et al. 2012; Zhuang et al. 2023). When people are exposed to the same social narrative, they often form similar interpretations, a characteristic that reflects the constraining influence of language, culture, and social norms (Nguyen et al. 2019). In neuroimaging, this shared understanding shows as similarities in brain activity across individuals, especially in higher-order brain regions involved in narrative comprehension and social cognition (Baek et al. 2022). During free recall, participants who share similar interpretations of animated narratives exhibit increased neural similarities in brain regions linked with higher-order cognition (Hasson et al. 2008; Nguyen et al. 2019).

Higher-order association networks, particularly the default mode network (DMN), play a central role in constructing shared social meaning by supporting mental simulation, perspective taking, and inference about others’ thoughts, intentions, and behaviors (Zatorre and Halpern 2005; Fox et al. 2015). Consistent with this framework, schizophrenia exhibit marked impairments in inferring others’ intentions, detecting socially relevant cues, and integrating contextual information into coherent social interpretations (Berretta et al. 2025). Previous neuroimaging studies have implicated abnormalities in the DMN and broader social-cognitive networks in schizophrenia (Hu et al. 2017; Romero-Garcia et al. 2020; Pan et al. 2023). However, most existing studies employ highly structured experimental paradigms, which feature explicit instructions and predefined correct answers, and focus on within-subject measures such as average activation levels or task performance—for example, selective attention tasks (Mannell et al. 2010) and semantic priming tasks (Jeong and Kubicki 2010). Although these approaches have provided important insights into specific cognitive processes, they offer limited understanding of whether patients and healthy individuals converge in their understanding and neural responses when processing continuous, naturalistic social information. Intersubject correlation (ISC), which quantifies the consistency of neural responses across individuals and provides a complementary approach for assessing psychological processes, including interpretation and emotional reactions (Nastase et al. 2019). Thus, ISC offers a unique opportunity to investigate whether the construction of shared social meaning during naturalistic social understanding is disrupted in schizophrenia.

Critically, social narratives are inherently ambiguous and require individuals to integrate prior beliefs with evolving contextual information to construct coherent interpretations (Nguyen et al. 2019). Individuals with schizophrenia show deficits in resolving ambiguous meanings and updating beliefs in response to new information, which may contribute to a greater reliance on pre-existing, often negative beliefs when interpreting ambiguous social cues (Ketteler et al. 2012; Savulich et al. 2015; Núñez et al. 2024). Paranoia represents a prominent manifestation of such altered social inference, characterized by a tendency to interpret ambiguous interpersonal situations as evidence of others’ hostile or threatening intentions (Horita, 2021; Pinkham et al., 2011; Sheffield et al., 2022). Consistent with this view, behavioral studies have demonstrated that higher levels of paranoia are associated with biased interpretations of ambiguous social situations (Raihani and Bell 2017; Barnby et al. 2020; Horita 2021). However, how individual differences in paranoia shape the organization of neural representations during complex social narrative processing in schizophrenia remains poorly understood. In order to address this unresolved question, it is necessary to move beyond global measures of neural synchrony and characterize the representational structure underlying individual differences. Intersubject correlation (ISC) quantifies the extent to which individuals exhibit similar neural responses while processing the same narrative (Nastase et al. 2019; Sonkusare et al. 2019; Finn et al. 2020), providing a measure of shared neural convergence during naturalistic cognition. However, ISC alone cannot reveal how clinical or psychological differences are reflected in the organization of neural representations across individuals. Intersubject Representational Similarity Analysis (IS-RSA) provides a complementary approach by testing whether the similarity of brain representations correspond to similarities in behavioral or clinical characteristics(Finn et al. 2020). Thus, IS-RSA provides a unique opportunity to determine whether paranoia-related differences are associated with altered neural representations during ambiguous social narrative processing.

To address these gaps, the present study adopts a naturalistic neuroimaging framework to investigate both shared and idiosyncratic neural representations during ambiguous social narrative processing in schizophrenia. By combining intersubject correlation (ISC) and intersubject representational similarity analysis (IS-RSA), we aimed to characterize two complementary aspects of social inference: the extent to which individuals converge on shared neural responses when interpreting the same social event, and how individual differences in paranoid ideation are reflected in the organization of neural representations. Patients with schizophrenia and matched healthy controls listened to an ambiguous social narrative during fMRI scanning and completed the Revised Green et al. Paranoid Thoughts Scale (R-GPTS). We hypothesized that schizophrenia would be associated with reduced neural convergence during social narrative processing, particularly within brain systems supporting social inference and contextual integration. Furthermore, we hypothesized that schizophrenia would be characterized by altered associations between paranoia and neural representations, reflecting a disrupted mapping between paranoid beliefs and the neural processes underlying the interpretation of ambiguous social information.

## Methods

### Participants

Patients with schizophrenia were recruited from the Second Mental Health Center of Beibei District of Chongqing. Healthy control participants were recruited from the local community in Chongqing. Participants who failed to complete the experiment or were excluded because of excessive head motion (see below) were excluded from subsequent analyses. The final sample consisted of 38 patients with schizophrenia (23 males, 15 females) and 46 healthy controls (20 males, 26 females) (Table 1). Inclusion criteria for patients with schizophrenia were: (1) age between 18 and 40 years; (2) meeting the diagnostic criteria for schizophrenia according to the International Classification of Diseases, Tenth Revision (ICD-10), with diagnosis confirmed by at least two qualified psychiatrists; (3) receiving stable doses of antipsychotic medication and being relatively symptomatically stable; (4) education level of junior middle school or above, with ability to understand story content and task requirements. Exclusion criteria for patients were: comorbid other severe mental disorders (e.g., bipolar disorder, major depressive disorder), neurological diseases (e.g., epilepsy, traumatic brain injury), intellectual disability or neurodevelopmental disorders; history of alcohol or substance abuse within the past 6 months; contraindications for MRI (e.g., metal implants, claustrophobia); severe physical illness, or recent severe self-injury, aggressive behavior, or acute psychotic symptom fluctuations precluding cooperation. Inclusion criteria for healthy controls were: (1) no history of psychiatric disorders; (2) no first-degree relatives diagnosed with mental disorders; (3) no history of neurological diseases; (4) no history of alcohol or substance abuse; (5) age between 18 and 40 years; (6) Healthy controls were matched to the schizophrenia group on age, sex, and years of education. All participants provided written informed consent prior to participation, and the study was approved by the Ethics Committee of Southwest University (approval number H25055). See table 1 for detailed demographic information.

**Table 1.** Participant Characteristics.

| | SZ(n=38) | HC(n=46) | $\chi^2$ | *p* |
| --- | --- | --- | --- | --- |
| Sex(female/male) | 15/23 | 26/20 | 2.421 | 0.12 |
| Age(years) | 32.84 ± 6.180 | 33.3 ± 7.257 | 0.310 | 0.757 |
| PANSS |  |  |  |  |
| total (n=34) | 56.88 ± 18.53 | - | - | - |
| positive (n=34) | 12.18 ± 4.23 | - | - | - |
| negative (n=34) | 15.47 ± 5.84 | - | - | - |
| general (n=34) | 29.24 ± 10.48 | - | - | - |
| Disease duration (years) | 12.12 ± 4.95 | - | - | - |
| Chlorpromazine equivalents (mg/day) | 507.5 ± 326.3 | - | - | - |

Notes: Four SZ patients had missing PANSS data, Disease duration and Chlorpromazine equivalents (mg/day); therefore, these three related analyses were based on 34 patients. Data are expressed as mean ± SD(SD: standard deviation). Abbreviations: PANSS, Positive and Negative Syndrome Scale.

### Stimulus

The narrative stimulus used in the present study was adapted from the original story materials developed by Emily S. Finn (Finn et al. 2018) to better suit the target population and experimental context. The revised narrative follows a series of uncertain events encountered by Dr. Wang Meng during a medical assistance mission in a remote mountain village, aiming to maintain participants’ engagement in narrative comprehension throughout the task (see Supplementary). To ensure consistency in language presentation and minimize potential confounding effects related to speaker variability, the story was narrated using an AI-generated Mandarin male voice. The final duration of the audio recording was 10min and 26s. During MRI scanning, participants listened to the audio recording of the story through headphones. Participants were instructed to remain attentive throughout the presentation and were not required to make any button responses or behavioral judgments during the task. Following the scanning session, participants completed a single-choice questionnaire regarding the story content to assess their level of attention and task engagement.

### Questionnaires

During a behavioral assessment session prior to scanning, participants completed the Revised Green et al. Paranoid Thoughts Scale (R-GPTS) (Green et al. 2008; Schlier et al. 2024). The R-GPTS was selected because it effectively assesses trait-level paranoia in clinical populations and, more importantly, is also applicable to subclinical and healthy populations (Finn et al. 2018). The full R-GPTS consists of two subscales, A and B, which assess ideas of persecution and reference, respectively(Schlier et al. 2024).

### MRI Data Acquisition

All MRI data were acquired at the Brain Imaging Center of Southwest University using a 3.0 T Siemens Prisma_fit scanner (syngo MR E11) equipped with a 64-channel head– neck coil. Participants were positioned supine with the head first (HFS). Structural images were acquired using a three-dimensional T1-weighted MPRAGE sequence (T1ISO_MPR, TFL+IR). Acquisition parameters were as follows: repetition time (TR) = 2.3 ms, echo time (TE) = 2.07 ms, inversion time (TI) = 1.1 ms, flip angle = 7°, slice thickness = 0.8 mm (isotropic high-resolution), base matrix = 320, reconstructed matrix = 280, and parallel acceleration factor = 2. These images were used for detailed anatomical analysis and co-registration of functional images.Functional images were collected using a gradient-echo echo-planar imaging (EPI) sequence (ep2d_bold) for task-based or resting-state BOLD acquisition. The acquisition parameters were: TR = 800 ms, TE = 30 ms, flip angle = 52°, slice thickness = 2.4 mm, matrix size = 90 × 90, multiband acceleration factor = 6, phase encoding direction = j−, effective echo spacing = 0.51 ms, total readout time = 45.391 ms, and pixel bandwidth = 2780 Hz/Px, yielding a high temporal resolution of 0.8 s.A single-band reference (SBRef) image with the same spatial resolution as the BOLD images was also acquired to improve functional image registration and signal calibration. The SBRef parameters were identical to the BOLD acquisition except for TR = 4.05 s.

To correct for susceptibility-induced distortions in EPI images, spin-echo EPI field map images (cmrr_mbep2d_se) were additionally acquired with the following parameters: TR = 7.04 s, TE = 80 ms, slice thickness = 2.4 mm, matrix size = 90 × 90, and total readout time = 45.391 ms. Two images with opposite phase-encoding directions—AP (j−) and PA (j)—were collected (blip-up/blip-down) to enable estimation of the susceptibility distortion field using the topup method and subsequent distortion correction of the BOLD images.

### Image Preprocessing

All imaging data were preprocessed using fMRIPrep 24.1.1 (Esteban et al. 2019). Susceptibility-induced distortions were corrected by estimating the B0 field inhomogeneity from the paired reverse phase-encoding (AP/PA) spin-echo EPI images using FSL TOPUP, and applying the resulting deformation field to the functional data. For anatomical preprocessing, T1-weighted (T1w) images underwent intensity non-uniformity correction using N4BiasFieldCorrection (ANTs) followed by skull stripping. Tissue segmentation into gray matter (GM), white matter (WM), and cerebrospinal fluid (CSF) was performed using FSL FAST. The T1w images were then nonlinearly normalized with ANTs to the MNI152NLin6Asym standard space.

Functional preprocessing included generation of a reference volume and head-motion correction using FSL MCFLIRT, followed by susceptibility distortion correction using the TOPUP-derived field. The distortion-corrected BOLD reference was rigidly co-registered to the individual T1w image using boundary-based registration (BBR) with 6 degrees of freedom. The motion correction, distortion correction, BOLD-to-T1w co-registration, and T1w-to-MNI nonlinear normalization transforms were concatenated and applied in a single interpolation step to resample the BOLD time series into standard MNI152NLin6Asym space, thereby achieving spatial normalization. During preprocessing, multiple confound regressors were computed, including framewise displacement (FD) and motion parameters. Volumes with excessive motion were flagged as outliers (FD > 0.5 mm). For subsequent denoising, we further regressed out WM and CSF signals and Friston’s 24-parameter motion model, and applied high-pass filtering to retain frequencies above 0.005 Hz, thereby reducing low-frequency drift. Participants were excluded if mean FD exceeded 0.3 mm or if more than 20% of volumes exceeded the FD threshold, resulting in the exclusion of two subjects.

### Intersubject Correlation Analysis

To examine whether schizophrenia affects shared neural responses during naturalistic social narrative processing, we employed intersubject correlation (ISC) analysis to quantify the similarity of neural activity across individuals (Nastase et al. 2019; Finn et al. 2020; Chen et al. 2020). First, whole-brain functional parcellation was conducted using the Schaefer-400 cortical atlas (Schaefer et al. 2018) combined with 32 subcortical regions (Tian et al. 2020) and 32 cerebellar regions (Diedrichsen et al. 2009),resulting in a total of 464 regions of interest (ROIs). Time series were then extracted for each participant during auditory story presentation.

Subsequently, within each group (patients with schizophrenia and healthy controls), pairwise correlations of the time series were computed across participants for each ROI, followed by Fisher’s z-transformation, yielding a participant-by-participant correlation matrix for each ROI. To compare the spatial distribution patterns of ISC between groups, the lower triangle of each ROI correlation matrix was averaged to obtain an ISC value for each participant. Group differences were assessed using a permutation test based on differences in mean ISC values. Specifically, group labels were randomly shuffled, and the mean ISC difference between groups was recalculated 10,000 times to construct a null distribution. P values were determined according to the position of the observed group mean difference within the null distribution, and false discovery rate (FDR) correction was applied across all ROIs using the Benjamini–Hochberg procedure (q < 0.05).

### Intersubject Representational Similarity Analysis

To further investigate how individual differences in paranoid ideation are reflected in neural representations during naturalistic social narrative processing, we employed intersubject representational similarity analysis (IS-RSA). Unlike ISC, which quantifies the convergence of neural responses across individuals exposed to the same stimulus, IS-RSA incorporates interindividual differences in behavioral or clinical characteristics to examine whether similarities in neural representations correspond to similarities in psychological traits (Finn et al. 2020; Chen et al. 2020). Specifically, IS-RSA was used to identify brain regions where neural similarity was associated with similarity in paranoid ideation across participants. First, behavioral similarity between all pairs of participants was calculated for each paranoia dimension, including “Totalscore”, “Persecution” and “Reference.” Based on the Anna Karenina model, it was assumed that individuals with higher behavioral scores would exhibit greater similarity in neural activity patterns, whereas individuals with lower scores would show greater variability. Therefore, the minimum score of each participant pair for a given dimension was used to construct the behavioral similarity matrix (Finn et al. 2020). To avoid the influence of differences in score ranges across paranoia dimensions on similarity estimation, all R-GPTS dimension scores were normalized to the range [0, 1] prior to constructing the behavioral similarity matrices. Subsequently, for each ROI, Spearman rank correlations were computed between the lower triangles of the intersubject neural similarity matrix and the behavioral similarity matrix, generating a brain–behavior matrix of “ISC region × paranoia dimension.” The statistical significance of these correlations was evaluated using the Mantel permutation test (Mantel 1967). Specifically, the rows and columns of either the neural or behavioral participant-by-participant similarity matrix were simultaneously randomized, and the Spearman correlation between the two matrices was recalculated. This procedure was repeated 1000 times to generate a null distribution of correlation coefficients, and p values were determined according to the position of the observed correlation coefficient within the null distribution. The p values of the two groups were respectively subjected to Benjamini-Hochberg FDR correction (q < 0.05). Finally, to examine whether the brain–behavior mapping relationship differed between patients with schizophrenia and healthy controls, group differences in IS-RSA correlation coefficients were compared. Specifically, differences in IS-RSA correlation coefficients between healthy controls and patients with schizophrenia were calculated for each ROI and paranoia dimension. Statistical significance was assessed using permutation testing by randomly shuffling group labels and recomputing the IS-RSA procedure 1000 times to construct a null distribution. Two-tailed p values were then calculated based on the position of the observed correlation coefficient differences within the null distribution. For both within-group IS-RSA results and between-group differences in IS-RSA, p values across all tested ROIs were corrected for multiple comparisons using the Benjamini–Hochberg false discovery rate (FDR) procedure.

## Results

### Reduced Neural Convergence During Ambiguous Social Narrative Processing in Schizophrenia

Story listening elicited robust intersubject synchrony across participants, indicating the emergence of shared neural responses during naturalistic social narrative processing. In both groups, the strongest ISC was observed in the primary auditory cortex and language-related regions along the superior temporal lobe. Importantly, significant synchrony extended beyond basic perceptual and linguistic systems to higher-order association regions, including frontal, parietal, and temporal cortices, as well as the posterior cerebellum, suggesting engagement of distributed networks supporting narrative comprehension and social meaning construction (Fig. 1a–b).

**Figure 1.**
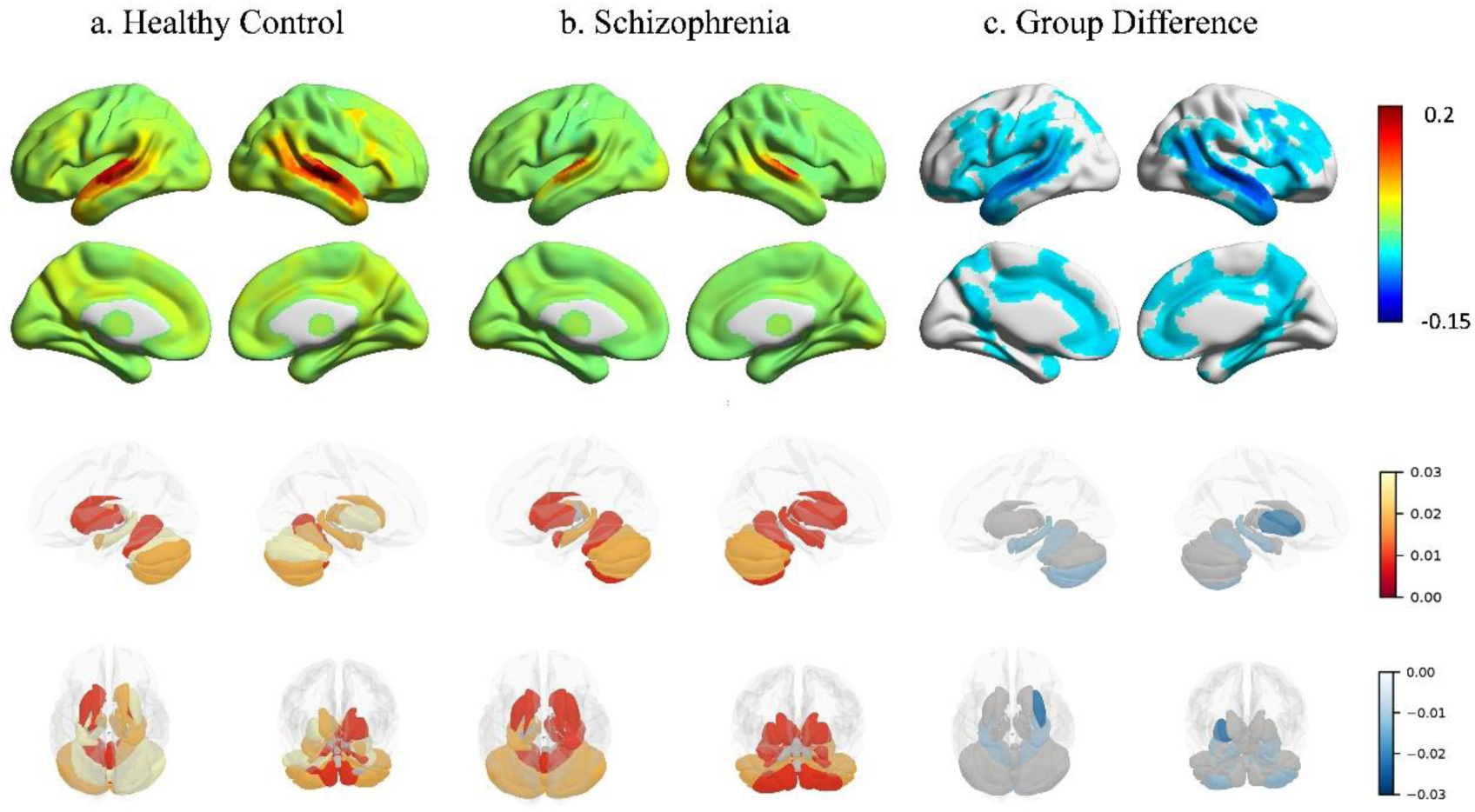
Group comparison of intersubject correlations, including the cortex, subcortex and cerebellum. a-b. In both groups, the strongest ISC was observed in the primary auditory cortex and language-related regions of the superior temporal gyrus, as well as in the frontal, parietal, and temporal cortical areas and the posterior cerebellum. c. Patients with schizophrenia showed decreased inter-individual synchrony in the temporal-parietal region (including the posterior middle temporal gyrus, angular gyrus, and inferior parietal cortex), bilateral insular regions, sensory-motor areas, and the left inferior frontal gyrus.

Compared with healthy controls, patients with schizophrenia exhibited reduced neural convergence during ambiguous social narrative processing Reduced intersubject synchrony was observed across distributed regions including temporoparietal areas (the posterior middle temporal gyrus, angular gyrus, and inferior parietal cortex), bilateral insula, sensorimotor regions, and left inferior frontal areas (Fig. 1c and Table S1). These regions primarily encompassed systems implicated in social inference, contextual integration, and salience attribution (Ilinsky et al. 1985; Van Overwalle 2009; Manoliu et al. 2014).

From the perspective of large-scale functional network organization (based on the Yeo’s seven-network), brain regions showing significantly reduced ISC were primarily distributed within the Default Mode Network, with additional involvement of the Visual and Control/Frontoparietal Networks.

### Reorganized Paranoia-Related Neural Representations in Schizophrenia

To examine whether paranoia-related individual differences were reflected in neural representations during ambiguous social narrative processing, we conducted intersubject representational similarity analysis (IS-RSA) separately in patients with schizophrenia and healthy controls. In healthy controls, higher total paranoia scores were associated with reduced neural similarity across a distributed network primarily involving the temporoparietal junction and adjacent temporal–parietal association regions, including the superior and middle temporal gyri, angular gyrus, and inferior parietal cortex. Additional associations were observed in frontal regions, including bilateral inferior and superior frontal gyri and the right middle frontal gyrus, as well as the right insula, limbic/paralimbic regions, posterior cerebellum, and left putamen (Fig. 2a).

**Figure 2.**
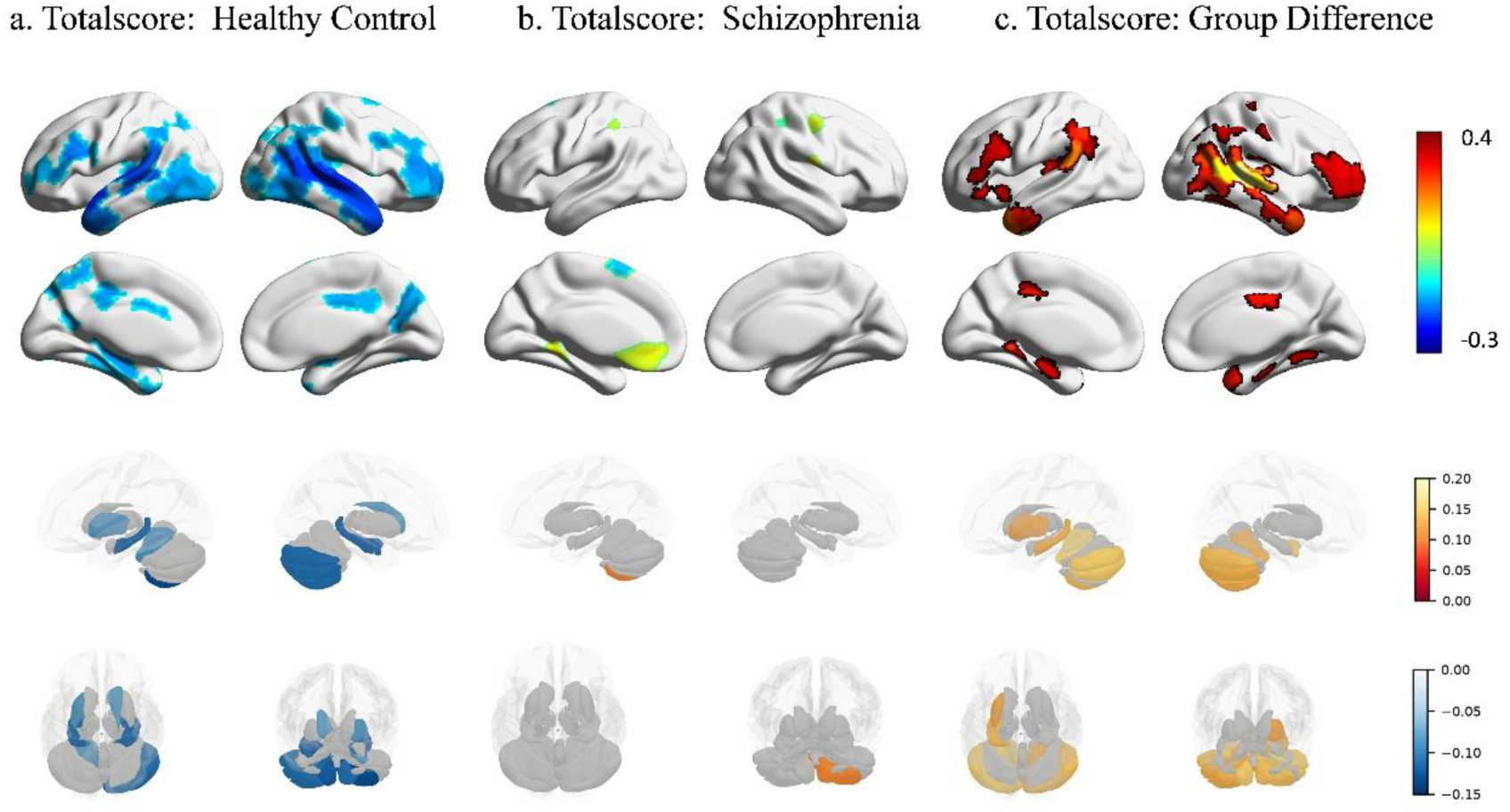
Group comparison of intersubject representational similarity analysis (IS-RSA) of the total score of Paranoid Thoughts Scale. a. In the healthy control group, the neural convergence decreased in the temporoparietal junction area, including the superior and middle temporal gyri, angular gyrus, and inferior parietal cortex. As well as the prefrontal area, limbic system/limbic paralimbic system, posterior lobe of the cerebellum and left putamen. b. In schizophrenia, the main involved areas include the bilateral inferior frontal gyrus, sensorimotor area, supplementary motor area, left medial orbitofrontal cortex and ACC. c. The inter-group comparison revealed that the paranoid-related IS-RSA values of schizophrenia patients were higher in the temporal-parietal junction, frontal cortex, insula, limbic-limbic band regions, posterior cerebellum and left putamen compared to the healthy control group.

In patients with schizophrenia, associations between paranoia and neural representations were more sparse, involving mainly frontoparietal motor-related regions and limbic–paralimbic areas, including bilateral inferior parietal lobules, sensorimotor regions, supplementary motor areas, left medial orbitofrontal cortex, and anterior cingulate cortex (ACC) (Fig. 2b). Direct group comparisons revealed higher paranoia-related IS-RSA values in schizophrenia relative to healthy controls across a distributed social-affective network, with prominent effects in the temporoparietal junction, frontal cortex, insula, limbic/paralimbic regions, posterior cerebellum, and left putamen (Fig. 2c and Table S2).

From the perspective of large-scale functional network organization (based on the Yeo’s seven-network), significant regions in the Totalscore dimension were predominantly located within the Control Network and the Default Mode Network, with additional involvement of the Visual, Somatomotor, and Limbic networks.

Although group differences in the Persecution and Reference dimensions largely overlapped with those observed for total paranoia, dimension-specific patterns emerged. Both dimensions showed higher IS-RSA values in schizophrenia across regions centered on the temporoparietal junction and extending to frontal cortex, insula, cerebellum, and basal ganglia. Persecution-related differences showed greater frontal involvement and additional reductions in occipital regions, whereas reference-related differences showed more prominent alterations in limbic regions and multiple thalamic subregions, with comparatively limited frontal involvement (Fig. 3 and Tables S3–S4).

**Figure 3.**
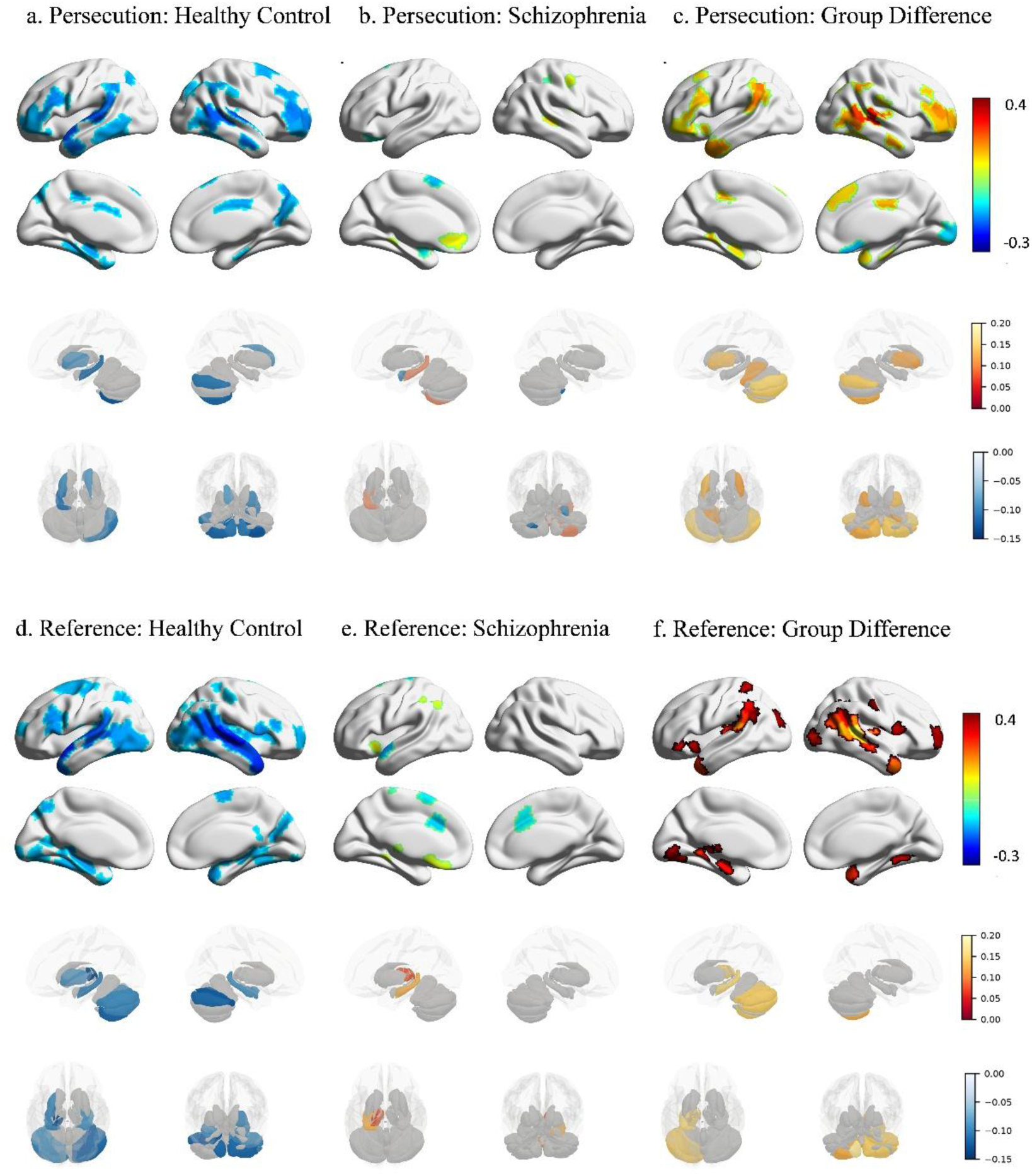
Group comparison of intersubject representational similarity analysis (IS-RSA) of the persecution and reference dimension, including the cortex, subcortex and cerebellum. a-c. Persecution dimension showed greater frontal involvement and additional reductions in occipital regions. d-f. Reference dimension showed more prominent alterations in limbic regions and multiple thalamic subregions.

## Discussion

Using naturalistic fMRI with an ambiguous social narrative, we investigated whether schizophrenia is associated with altered convergence of neural responses during ecologically valid social information processing and whether such alterations are linked to individual differences in paranoid ideation. Two main findings emerged. First, patients with schizophrenia showed significantly reduced intersubject synchrony relative to healthy controls during social narrative processing, particularly within distributed systems supporting social inference, contextual integration, and salience processing, including temporoparietal, temporal, insular, and inferior frontal regions. Second, intersubject representational similarity analysis revealed altered associations between paranoid ideation and neural representations, suggesting that schizophrenia involves not only reduced convergence in processing shared social information but also a reorganization of paranoia-related representations during ambiguous interpersonal processing. Together, these findings demonstrate that schizophrenia disrupts both shared neural alignment during social meaning construction and the organization of symptom-relevant neural representations.

Methodologically, while traditional paradigms typically employ brief, structured stimuli and explicit task instruction, this study pushes beyond conventional approaches in schizophrenia social cognition research by combining naturalistic fMRI with ambiguous social narratives, enabling the capture of ecologically valid, continuous social information processing (Finn et al. 2018; Evans et al. 2020; Gorrino et al. 2024; Cavieres et al. 2025). By combining ISC to measure shared neural convergence and IS-RSA to assess individual differences in paranoia-related representational structure (Nastase et al. 2019; Sonkusare et al. 2019; Finn et al. 2020), the present design reveals both group-level disruptions in social meaning construction and symptom-relevant variability in neural processing. This framework highlights a key advantage of the current study: it bridges ecological social cognition with clinically relevant neural representations, providing a more nuanced understanding of how schizophrenia affects the processing of ambiguous social information.

Reduced intersubject synchrony in schizophrenia may reflect disrupted neural convergence underlying the construction of shared social meaning during naturalistic social experiences(Wilson et al. 2008; Jääskeläinen et al. 2020; Finn et al. 2020). Although both groups exhibited robust synchronization in auditory and language-related cortices, patients showed significantly reduced ISC in the temporoparietal junction, temporal cortex, insula, and inferior frontal gyrus, suggesting that the observed group differences are unlikely to be explained simply by impaired sensory processing or failure to follow the narrative. Rather, these regions collectively constitute a distributed network supporting contextual integration, mental-state inference, belief updating, and social salience processing, all of which are fundamental to successful social understanding (Sankar et al. 2023; Blain et al. 2023; Zhang et al. 2025; Campos-Sousa and Almeida 2026). Healthy individuals tend to generate similar neural responses because they construct comparable internal models of characters’ intentions and unfolding social events(Baek et al. 2022; Khajehnejad et al. 2025). In contrast, impairments in mentalizing, contextual integration, and flexible belief updating in schizophrenia may hinder the convergence of these internal models (Standke et al. 2021; Gibbs-Dean et al. 2023), leading to increasingly idiosyncratic interpretations of the same ambiguous social narrative (Mennen et al. 2022). Consistent with this account, reduced ISC was predominantly distributed within the default mode, control/frontoparietal, and visual networks, indicating that schizophrenia disrupts the coordinated large-scale neural architecture supporting context-dependent social inference rather than isolated cognitive processes. Together, these findings suggest that reduced ISC reflects diminished convergence of internal representations of others’ actions, intentions, and emotional states across patients, providing a neural basis for impaired shared social understanding during naturalistic social cognition in schizophrenia.

Beyond reduced intersubject neural synchrony, the IS-RSA findings further demonstrate that the relationship between paranoid beliefs and neural representational organization is fundamentally altered during the processing of ambiguous social narratives. In healthy controls, higher levels of paranoid ideation were associated with lower neural representational similarity across brain regions involved in social cognition, salience processing, affective evaluation, and action prediction (Blain et al. 2023; Sobral et al. 2025), suggesting that even subclinical variations in paranoia may reduce convergence toward normative interpretations of social events. In contrast, patients with schizophrenia exhibited significantly stronger associations between paranoia similarity and neural representational similarity across a distributed network encompassing temporoparietal, frontal, insular, limbic, and cerebellar regions. Rather than reflecting greater homogeneity of neural responses, this pattern indicates that social representations become increasingly organized by pre-existing paranoid beliefs. In healthy individuals, interpretations of ambiguous social situations are primarily constrained by contextual information, leading to relatively convergent neural representations across individuals (Nguyen et al. 2019; Baek and Parkinson 2022). In contrast, in patients with schizophrenia, impairments in contextual integration and theory of mind may weaken reliance on external social cues, and under the influence of paranoid ideation, result in the attribution of internally generated thoughts to perceived external threats (Kapur 2003; Ricarte et al. 2017; Wu et al. 2023).

Importantly, dimensional analyses further revealed both shared and distinct neural signatures of persecution and reference ideation. Both dimensions exhibited representational reorganization centered on temporoparietal, temporal, and insular regions, supporting the notion that different forms of paranoia share a common disruption in the neural architecture underlying social meaning construction. However, persecution was additionally associated with more extensive frontal involvement together with reduced representational similarity in occipital regions, suggesting greater engagement of belief evaluation, cognitive control, and threat-related interpretation during social inference (Yang et al. 2015; Wu et al. 2023). By contrast, reference ideation showed relatively greater involvement of limbic structures and thalamic nuclei, regions implicated in self-referential processing, associative memory, and salience integration (Helpman et al. 2017). These findings suggest that although different dimensions of paranoia are grounded in a common social-cognitive representational framework, they bias social interpretation through partially distinct representational mechanisms. Taken together, the IS-RSA findings extend the ISC results by demonstrating that schizophrenia is characterized not only by reduced neural convergence during social narrative processing but also by a reorganization of the mapping between paranoid beliefs and the neural systems supporting social meaning construction.

Several limitations should be considered when interpreting these findings. First, patients were clinically stable and receiving antipsychotic medication, making it difficult to disentangle illness-related alterations from medication or chronicity effects; replication in first-episode, unmedicated, or longitudinal cohorts would help clarify whether disrupted neural synchrony and altered paranoia-related representations reflect core features of psychosis. Second, although the post-scan questionnaire confirmed general task engagement, the study did not collect detailed moment-to-moment behavioral measures of narrative interpretation, perceived threat, or character intention. This was partly an intentional design choice, as introducing continuous in-scanner ratings would impose additional task demands and potentially alter the spontaneous engagement with the narrative, which is a key feature that naturalistic paradigms aim to preserve (Nau et al. 2024). Future studies combining naturalistic stimuli with post hoc or carefully designed behavioral assessments may further clarify the relationship between neural convergence, subjective interpretation, and paranoid ideation. Third, the use of a single ambiguous narrative limits conclusions about stimulus generalizability, as effects may depend on story content; future work should test multiple narratives varying in social ambiguity, emotional salience, and threat content (Finn et al. 2018).

In conclusion, this study demonstrates that schizophrenia is characterized by impaired neural convergence during ambiguous social narrative processing and a reorganization of paranoia-related neural representations. These findings suggest that schizophrenia disrupts the ability to construct shared social meaning from dynamic interpersonal information, while paranoid ideation further alters how ambiguous social cues are interpreted and represented within distributed social-cognitive and affective systems. By integrating naturalistic fMRI with intersubject analyses, this study provides an ecologically grounded framework for understanding how disruptions in shared neural alignment and idiosyncratic representational organization contribute to abnormal social inference and paranoia in schizophrenia.

## Data Availability

All data generated in this study are available from the authors upon reasonable request.

## Supplementary material

Supplementary material is available at Psychoradiology Journal online.

## Acknowledgements

This work was supported by the National Natural Science Foundation of China (Grant Nos. 32300861 and 82202247) and the Natural Science Foundation of Chongqing (CSTB2023NSCQ-MSX0896).

## Reference

Baek EC, Hyon R, López K, et al (2022) In-degree centrality in a social network is linked to coordinated neural activity. Nat Commun 13:1118. 10.1038/s41467-022-28432-3

Baek EC, Parkinson C (2022) Shared understanding and social connection: Integrating approaches from social psychology, social network analysis, and neuroscience. Soc Personal Psychol Compass 16: e12710. 10.1111/spc3.12710

Barnby JM, Deeley Q, Robinson O, et al (2020) Paranoia, sensitization and social inference: Findings from two large-scale, multi-round behavioural experiments. R Soc Open Sci 7: 191525. 10.1098/rsos.191525

Berretta SA, Oliver LD, Hyatt CS, et al (2025) Domain-specific associations between social cognition and aggression in schizophrenia spectrum disorders. Schizophr Res Cogn 41:100361. 10.1016/j.scog.2025.100361

Blain SD, Taylor SF, Rutherford SE, et al (2023) Neurobehavioral Indices of Gaze Perception Are Associated With Social Cognition Across Schizophrenia Patients and Healthy Controls. Journal of Psychopathology and Clinical Science 132:733–748. 10.1037/abn0000846

Campos-Sousa RN, Almeida KJ (2026) Neurology of cognition and social behavior. A narrative review of neurobiological bases and clinical aspects. Dement Neuropsychol 20: e20250301. 10.1590/1980-5764-dn-2025-0301

Cavieres A, Acuña V, Arancibia M, Escobar C (2025) Advances in the ecological validity of research on social cognition in schizophrenia: A systematic review of the literature. Schizophr Res Cogn 39:100333. 10.1016/j.scog.2024.100333

Chen PHA, Jolly E, Cheong JH, Chang LJ (2020) Intersubject representational similarity analysis reveals individual variations in affective experience when watching erotic movies. Neuroimage 216: 116851. 10.1016/j.neuroimage.2020.116851

Diedrichsen J, Balsters JH, Flavell J, et al (2009) A probabilistic MR atlas of the human cerebellum. Neuroimage 46:39–46. 10.1016/j.neuroimage.2009.01.045

Esteban O, Markiewicz CJ, Blair RW, et al (2019) fMRIPrep: a robust preprocessing pipeline for functional MRI. Nat Methods 16:111–116. 10.1038/s41592-018-0235-4

Evans TC, Bar-Haim Y, Fox NA, et al (2020) Neural mechanisms underlying heterogeneous expression of threat-related attention in social anxiety. Behaviour Research and Therapy 132: 103657. 10.1016/j.brat.2020.103657

Finn ES, Corlett PR, Chen G, et al (2018) Trait paranoia shapes inter-subject synchrony in brain activity during an ambiguous social narrative. Nat Commun 9: 2043. 10.1038/s41467-018-04387-2

Finn ES, Glerean E, Khojandi AY, et al (2020) Idiosynchrony: From shared responses to individual differences during naturalistic neuroimaging. Neuroimage 215: 116828. 10.1016/j.neuroimage.2020.116828

Fox KCR, Spreng RN, Ellamil M, et al (2015) The wandering brain: Meta-analysis of functional neuroimaging studies of mind-wandering and related spontaneous thought processes. Neuroimage 111:611–621. 10.1016/j.neuroimage.2015.02.039

Gibbs-Dean T, Katthagen T, Tsenkova I, et al (2023) Belief updating in psychosis, depression and anxiety disorders: A systematic review across computational modelling approaches. Neurosci Biobehav Rev 147:105087. 10.1016/j.neubiorev.2023.105087

Gorrino I, Rossetti MG, Girelli F, et al (2024) A critical overview of emotion processing assessment in non-Affective and affective psychoses. Epidemiol Psychiatr Sci 33:1–5. 10.1017/S204579602400009X

Green CEL, Freeman D, Kuipers E, et al (2008) Measuring ideas of persecution and social reference: the Green et al. Paranoid Thought Scales (GPTS). Psychol Med 38:101–111. 10.1017/S0033291707001638

Hasson U, Ghazanfar AA, Galantucci B, et al (2012) Brain-to-brain coupling: a mechanism for creating and sharing a social world. Trends Cogn Sci 16:114–121. 10.1016/j.tics.2011.12.007

Hasson U, Yang E, Vallines I, et al (2008) A Hierarchy of Temporal Receptive Windows in Human Cortex. The Journal of Neuroscience 28:2539–2550. 10.1523/JNEUROSCI.5487-07.2008

Helpman L, Zhu X, Suarez-Jimenez B, et al (2017) Sex Differences in Trauma-Related Psychopathology: a Critical Review of Neuroimaging Literature (2014– 2017). Curr Psychiatry Rep 19:104. 10.1007/s11920-017-0854-y

Horita Y (2021) Conjecturing Harmful Intent and Preemptive Strike in Paranoia. Front Psychol 12: 726081. 10.3389/fpsyg.2021.726081

Hu M-L, Zong X-F, Mann JJ, et al (2017) A Review of the Functional and Anatomical Default Mode Network in Schizophrenia. Neurosci Bull 33:73–84. 10.1007/s12264-016-0090-1

Ilinsky IA, Jouandet ML, Goldman-Rakic PS (1985) Organization of the nigrothalamocortical system in the rhesus monkey. Journal of Comparative Neurology 236:315–330. 10.1002/cne.902360304

Jääskeläinen IP, Klucharev V, Panidi K, Shestakova AN (2020) Neural Processing of Narratives: From Individual Processing to Viral Propagation. Front Hum Neurosci 14: 253. 10.3389/fnhum.2020.00253

Jeong B, Kubicki M (2010) Reduced task-related suppression during semantic repetition priming in schizophrenia. Psychiatry Res Neuroimaging 181:114–120. 10.1016/j.pscychresns.2009.09.005

Kapur S (2003) Psychosis as a State of Aberrant Salience: A Framework Linking Biology, Phenomenology, and Pharmacology in Schizophrenia. American Journal of Psychiatry 160:13–23. 10.1176/appi.ajp.160.1.13

Ketteler D, Theodoridou A, Ketteler S, Jäger M (2012) High Order Linguistic Features Such as Ambiguity Processing as Relevant Diagnostic Markers for Schizophrenia. Schizophr Res Treatment 2012:1–7. 10.1155/2012/825050

Khajehnejad M, Habibollahi F, Loeffler A, et al (2025) Dynamic Network Plasticity and Sample Efficiency in Biological Neural Cultures: A Comparative Study with Deep Reinforcement Learning. Cyborg and Bionic Systems 6:0336. 10.34133/cbsystems.0336

Kuhlmann M, Meuer J, Bening CR (2023) Interorganizational Sensemaking of the Transition Toward a Circular Value Chain. Organ Environ 36:411–441. 10.1177/10860266231162057

Maitlis S (2005) The Social Processes of Organizational Sensemaking. Academy of Management Journal 48:21–49. 10.5465/amj.2005.15993111

Mannell M V., Franco AR, Calhoun VD, et al (2010) Resting state and task-induced deactivation: A methodological comparison in patients with schizophrenia and healthy controls. Hum Brain Mapp 31:424–437. 10.1002/hbm.20876

Manoliu A, Riedl V, Zherdin A, et al (2014) Aberrant Dependence of Default Mode/Central Executive Network Interactions on Anterior Insular Salience Network Activity in Schizophrenia. Schizophr Bull 40:428–437. 10.1093/schbul/sbt037

Mantel N (1967) The detection of disease clustering and a generalized regression approach. Cancer Res 27:209–20

Mennen AC, Nastase SA, Yeshurun Y, et al (2022) Real-time neurofeedback to alter interpretations of a naturalistic narrative. Neuroimage: Reports 2: 100111. 10.1016/j.ynirp.2022.100111

Nastase SA, Gazzola V, Hasson U, Keysers C (2019) Measuring shared responses across subjects using intersubject correlation. Soc Cogn Affect Neurosci 14:669– 687. 10.1093/scan/nsz037

Nau M, Schmid AC, Kaplan SM, et al (2024) Centering cognitive neuroscience on task demands and generalization. Nat Neurosci 27:1656–1667. 10.1038/s41593-024-01711-6

Nguyen M, Vanderwal T, Hasson U (2019) Shared understanding of narratives is correlated with shared neural responses. Neuroimage 184:161–170. 10.1016/j.neuroimage.2018.09.010

Núñez D, Rodríguez-Delgado J, Castillo RD, et al (2024) Effect of prior beliefs and cognitive deficits on learning in first-episode schizophrenia patients. Schizophr Res Cogn 38: 100318. 10.1016/j.scog.2024.100318

Pan Y, Wen Y, Wang Y, et al (2023) Interpersonal coordination in schizophrenia: a concise update on paradigms, computations, and neuroimaging findings. Psychoradiology 3:1–5. 10.1093/psyrad/kkad002

Pinkham AE, Brensinger C, Kohler C, et al (2011) Actively paranoid patients with schizophrenia over attribute anger to neutral faces. Schizophr Res 125:174–178. 10.1016/j.schres.2010.11.006

Raihani NJ, Bell V (2017) Paranoia and the social representation of others: A large-scale game theory approach. Sci Rep 7: 4544. 10.1038/s41598-017-04805-3

Ricarte JJ, Ros L, Latorre JM, Watkins E (2017) Mapping autobiographical memory in schizophrenia: Clinical implications. Clin Psychol Rev 51:96–108. 10.1016/j.cpr.2016.11.004

Romero-Garcia R, Seidlitz J, Whitaker KJ, et al (2020) Schizotypy-Related Magnetization of Cortex in Healthy Adolescence Is Colocated With Expression of Schizophrenia-Related Genes. Biol Psychiatry 88:248–259. 10.1016/j.biopsych.2019.12.005

Sankar A, Shen X, Colic L, et al (2023) Predicting depressed and elevated mood symptomatology in bipolar disorder using brain functional connectomes. Psychol Med 53:6656–6665. 10.1017/S003329172300003X

Savulich G, Freeman D, Shergill S, Yiend J (2015) Interpretation Biases in Paranoia. Behav Ther 46:110–124. 10.1016/j.beth.2014.08.002

Schaefer A, Kong R, Gordon EM, et al (2018) Local-Global Parcellation of the Human Cerebral Cortex from Intrinsic Functional Connectivity MRI. Cerebral Cortex 28:3095–3114. 10.1093/cercor/bhx179

Schlier B, Lincoln TM, Kingston JL, et al (2024) Cross-cultural validation of the revised Green et al., paranoid thoughts scale. Psychol Med 54:1985–1991. 10.1017/S0033291724000072

Sheffield JM, Suthaharan P, Leptourgos P, Corlett PR (2022) Belief Updating and Paranoia in Individuals With Schizophrenia. Biol Psychiatry Cogn Neurosci Neuroimaging 7:1149–1157. 10.1016/j.bpsc.2022.03.013

Sobral M, Guiomar R, Rezaeian M, et al (2025) Neural correlates of peripartum depression: a systematic review, meta-analysis and comparison to major depressive disorder. Mol Psychiatry 30:5979–6006. 10.1038/s41380-025-03227-2

Sonkusare S, Breakspear M, Guo C (2019) Naturalistic Stimuli in Neuroscience: Critically Acclaimed. Trends Cogn Sci 23:699–714. 10.1016/j.tics.2019.05.004

Standke I, Trempler I, Dannlowski U, et al (2021) Cerebral and behavioral signs of impaired cognitive flexibility and stability in schizophrenia spectrum disorders. Neuroimage Clin 32: 102855. 10.1016/j.nicl.2021.102855

Tian Y, Margulies DS, Breakspear M, Zalesky A (2020) Topographic organization of the human subcortex unveiled with functional connectivity gradients. Nat Neurosci 23:1421–1432. 10.1038/s41593-020-00711-6

Van Overwalle F (2009) Social cognition and the brain: A meta-analysis. Hum Brain Mapp 30:829–858. 10.1002/hbm.20547

Wilson SM, Molnar-Szakacs I, Iacoboni M (2008) Beyond Superior Temporal Cortex: Intersubject Correlations in Narrative Speech Comprehension. Cerebral Cortex 18:230–242. 10.1093/cercor/bhm049

Wu Y, Song S, Shen Y (2023) Characteristics of theory of mind impairment and its relationship with clinical symptoms and neurocognition in patients with schizophrenia. BMC Psychiatry 23: 711. 10.1186/s12888-023-05224-7

Yang C, Zhang T, Li Z, et al (2015) The relationship between facial emotion recognition and executive functions in first-episode patients with schizophrenia and their siblings. BMC Psychiatry 15: 241. 10.1186/s12888-015-0618-3

Zatorre RJ, Halpern AR (2005) Mental Concerts: Musical Imagery and Auditory Cortex. Neuron 47:9–12. 10.1016/j.neuron.2005.06.013

Zhang H, Wang X, Chen G, et al (2025) Noninvasive Intracranial Source Signal Localization and Decoding with High Spatiotemporal Resolution. Cyborg and Bionic Systems 6:0206. 10.34133/cbsystems.0206

Zhuang T, Kabulska Z, Lingnau A (2023) The Representation of Observed Actions at the Subordinate, Basic, and Superordinate Level. The Journal of Neuroscience 43:8219–8230. 10.1523/JNEUROSCI.0700-22.2023

