## Supplementary material for "Reduced Neural Convergence and Reorganized Paranoia-Related Representations During Ambiguous Social Narrative Processing in Schizophrenia"

**Story**

**Chinese version**

冬日的午后，王萌医生在成都的医院办公室里整理病历，身心疲惫。一封邮件打破了她的宁静，标题是“医生，我需要你的援助”，发件人是李伟博士。王萌对这个名字既熟悉又陌生，邮件中提到，李伟博士在云南一个偏远山村的诊所工作，因紧急情况需返回北京，他希望王萌能去接替他的工作几个月。因为诊所是村民们唯一的医疗保障，若无人接替，公益组织将关闭它。王萌心中涌起一股冲动。

她在成都的生活虽安稳，但内心深处一直渴望一场冒险。经过一夜的思量，她决定接受这个挑战。

一周后，王萌从成都出发，经昆明中转，飞机抵达滇西边境的小城芒市。走出机场，山野的微风拂面而来，让她感到一种久违的清新。

在机场候机厅，一个叫张强的男人来接她去山村。他们乘坐一辆破旧的越野车，沿着崎岖的山路前行。窗外的景色不断变换，山间薄雾缭绕，村落星星点点。车子最终停在一条小河边，一辆马车正等候在那里，将载她踏上最后一程。

马车缓缓前行，王萌的心情也逐渐放松。然而，当马车停下，她发现自己置身于一片寂静之中，四周是连绵的青山和茂密的山林。马车夫离开后，她感到一丝不安。就在这时，一个男人提着油灯从树林中走出，他是村长老陈。

他热情地欢迎王萌，说全村都在盼着她的到来。老陈带着王萌穿过山林，来到打谷场的木屋。木屋里只摆放着一些简单的家具。老陈告诉她先休息，明天再好好聊聊。王萌疲惫地躺在床上，很快进入了梦乡。

第二天清晨，阳光透过木屋的缝隙洒在王萌脸上。她坐起身来，突然，角落里传来一阵声响，一个小女孩正低着头扫地。王萌吓了一跳，女孩抬起头，露出怯生生的眼神。王萌指了指自己，说：“我叫王萌。”女孩也指了指自己，轻声说：“我叫小梅。”

“老陈呢？”王萌比划着问道，小梅点点头，示意王萌跟着她出去。王萌跟着小梅走出木屋，发现老陈正站在村口，和几个穿着当地特色服装的人交谈。老陈注意到王萌，便对其他人说了几句，其他人也纷纷转头看向她。王萌一时间觉得不太自在。

“王萌医生，早上好啊！”老陈热情地说道，“我带你去诊所看看吧。”“好啊，有劳了。”王萌连忙应道，心里满是期待。

老陈带着王萌沿着一条蜿蜒的小路来到诊所。推门进去，只见里面两张简陋的木板床直接铺在地上，还有一张用木板拼凑而成的桌子。仅有的医疗设备就是一个旧式听诊器、一个体温计和一盒所剩无几的纱布。“这……李伟医生说诊所医疗物资挺齐全的呀？”王萌有些疑惑地说道。

“李伟医生？”老陈一脸困惑地重复道，“哦，对对，就是李伟。组织上答应给我们寄物资过来，可一直没到。您也知道，山里交通不便，物资运输总是容易耽搁。” “没事。”王萌说道。在新一批物资到来之前，她只能先依靠自己随身携带的小药箱维持诊疗。

小梅成了王萌的好伙伴。每天清晨，她都会来给王萌送早餐。晚上，王萌回到木屋，小梅会来帮忙打扫。小梅还会教王萌说当地方言，王萌也开始教小梅普通话。

王萌在诊所的第一周还算顺利。前来就诊的村民大多是一些小毛病。王萌感觉，很多人来并不是因为身体真的不舒服，而是好奇她这个外乡人。尽管如此，王萌并不介意，反而觉得他们都挺有趣的。大多数村民说的是当地方言，当老陈在身边时，他能帮忙翻译，但更多时候，王萌只能依靠手势和表情与他们交流。村民们都很感激她的帮助，王萌也很有成就感。

一周后，诊所来了一位名叫阿彩的老妇人，她发高烧，胸口和后背起了奇怪的红疹。王萌给她开了药，但病情并未好转。那天晚上，老陈和另一个村民来到诊所，站在门口，不愿靠近，仿佛害怕被传染。王萌问老陈村里是否有人出现过类似的症状，老陈摇头否认，但另一个人脸上却闪过一丝异样的神情。第二天，阿彩不见了，小梅说她被人带走了，但不愿多说。

第二个星期，村里举办了一场热闹的节日庆典，村民们载歌载舞，欢声笑语回荡在山谷之间。老陈坐在王萌身边，感激地说：“我们全村人都特别感激您不远千里来给我们看病，真不知道没有您我们该怎么办。”王萌心里暖暖的，为自己能参与这次冒险而感到高兴。

尽管在山村里过得挺开心，但王萌偶尔还是会想念家乡。一天清晨，她发现木屋后的小山坡有微弱信号，赶忙给闺密林林打电话，把山村里的所见所闻都讲给她听，还提到了那个奇怪发烧的病人。

挂断电话，王萌回木屋准备午休，敲门声响起。老陈匆匆进来：“村里发现盗伐者在禁伐区砍树，得赶紧联系林业部门。可卫星电话坏了，要借下你的手机。”王萌把手机和充电器递给他：“不着急，您慢慢用。”老陈离开后，诊所又来了个发烧的年轻人，他出红疹、咳得厉害。王萌陪他到深夜，可他还是去世了。尽管知道自己尽力了，但王萌还是悲痛愧疚。

几天后，王萌迎来了第三个奇怪发烧的病人，阿强，一个在村里颇有威望的族长，也是老陈的好友。幸运的是，阿强的病情并不严重，王萌用药物和草药为他治疗，最终他康复了。阿强的家人送来许多土特产表示感谢。王萌感到自己终于融入了这个新环境。

然而，王萌心里始终为那些因发烧而受苦的村民感到沮丧。她绞尽脑汁地思考可能有效的药物，还列了一张长长的药品清单给老陈。老陈虽然还没把手机还给她，但王萌也不想催促他。

第二天早上，王萌去诊所的路上，看到老陈正站在村子中央，和那几个族长模样的人交谈。当她走近时，男人们突然都停下了谈话，这让王萌觉得她可能无意间打扰了什么重要的事情。把药品清单交给老陈后，他告诉王萌他会尽快下单采购。可王萌突然想起老陈之前承诺的那批物资还没到。那天晚上，王萌躺在床上翻来覆去睡不着。她满脑子都在想，这会不会是一种从未见过的疾病？有时，王萌会觉得村民们似乎有些事情瞒着她。难道他们对这种疾病知道得比说出来的多？他们是想考验我的医术？或者觉得让自己生病，才是获得医疗帮助的唯一办法吗？但很快，王萌就为自己的这种想法感到羞愧。村民们对她热情友善，没有理由认为他们早在她来之前就知道发烧的事情，而且，李伟医生在邮件里也没有提及此事。

王萌在胡思乱想之际，突然意识到小梅那天晚上没来木屋找她。她猜测这小姑娘可能是在帮家里人准备迎接雨季，虽然很想念小梅的陪伴，但也只能作罢。

第二天早上吃早饭时，小梅依然没出现。王萌在诊所忙碌了一整天，晚上回到木屋，也没有等到小梅。王萌又担心又孤单，决定去小梅家看看。

王萌从未进过小梅家的木屋，虽然她知道是哪一间。她小心翼翼地穿过空地，避开雨水积成的水洼。来到小梅家木屋前，她先轻轻敲了敲门，屋内没有任何回应。她不想贸然侵犯别人的隐私，正要转身离开，可关心和好奇又让她停下了脚步，最终还是推开了门。

眼前的一幕让王萌大惊失色。小梅正躺在床上，不停咳嗽，身体还在艰难地挣扎。一个女人和一个男人正弯着腰，试图喂她喝下一碗汤药。小梅的上衣被脱掉，借着微弱的炉火光，王萌清楚地看到她胸前的红疹。王萌倒吸一口凉气，抬头一看，那个男人竟是老陈，他的眼睛里满是泪水。

“小梅是您的女儿？”王萌震惊地问道。她怎么能一点都不知道呢？老陈点了点头，眼神里满是绝望。

“我马上回来。”王萌说完，转身准备冲向诊所去拿仅剩的退烧药。可刚转身，她就瞥见手机正放在门边的桌子上，本能地抓起手机就往回跑。

王萌没有直接去诊所。她提着油灯，沿着通往河边的小路一路奔去。气喘吁吁地来到河边，她坐在一块石头上，呆呆地望着眼前混浊的河水。她多么希望能有船经过呀，可四周一片寂静。此刻，她才真正意识到自己身处如此偏远而又孤立无援的地方。

小梅到底病了多久了呢？她为什么一直瞒着自己？王萌突然想起小梅那双为她准备早餐、叠衣服、铺床的手，不禁怀疑这病会不会传染。要是自己已经被感染了怎么办？

王萌看着手里的手机，努力让自己冷静下来，可心里的焦虑和恐惧却像野火般蔓延。她急切地想逃离这里，远离可能的病魔，可一想到要抛下村民们，尤其是信任她的小梅，她又怎能忍心把他们丢在这样的困境之中？可若留下来，面对这种未知的疾病，她又不知自己是否有能力治愈，甚至还可能危及自己的生命。

她陷入了深深的纠结和痛苦之中，不知该如何抉择。

**English version**

On a winter afternoon, Dr. Wang Meng sat in her office at a hospital in Chengdu, sorting through medical records. Exhausted both physically and mentally, she longed for a quiet moment. Then an email appeared in her inbox, breaking the silence. The subject line read: “Doctor, I Need Your Help.” It was from Dr. Li Wei. The name felt both familiar and distant. In the email, Dr. Li explained that he had been working at a small clinic in a remote mountain village in Yunnan. An emergency required him to return to Beijing immediately, and he hoped Wang Meng could take over his position for a few months. The clinic was the villagers’ only source of medical care, and without a replacement, the nonprofit organization supporting it would be forced to shut it down. Something stirred inside her.

Life in Chengdu was comfortable and predictable, but deep down she had always yearned for an adventure. After a sleepless night of deliberation, she decided to accept the challenge.

A week later, Wang Meng left Chengdu, transferred in Kunming, and landed in Mangshi, a small city on Yunnan's western frontier.

As she stepped out of the airport, a cool breeze swept down from the surrounding mountains, carrying with it the fresh scent of the wilderness. It was a feeling she hadn’t experienced in years. Waiting for her in the arrival hall was a man named Zhang Qiang, who had come to take her to the village.

They climbed into a battered off-road vehicle and drove along winding mountain roads. Outside the window, the landscape shifted constantly—mist drifted through the valleys, and scattered villages dotted the mountainsides. Eventually, the vehicle stopped beside a narrow river. Waiting there was a horse-drawn cart, which would carry her the rest of the way.

As the cart creaked slowly along the trail, Wang Meng gradually began to relax. But when it finally came to a stop, she found herself surrounded by silence. Rolling green mountains stretched endlessly in every direction, cloaked in dense forest. After the driver departed, a faint sense of unease crept over her. Just then, a man carrying an oil lantern emerged from the trees.

He was Old Chen, the village chief.

He greeted Wang Meng warmly, telling her that everyone in the village had been eagerly awaiting her arrival. Leading her through the forest, he brought her to a wooden cabin beside the village threshing ground. The cabin was sparsely furnished, containing only a few simple pieces of furniture. “You should get some rest,” Old Chen said. “We'll have plenty of time to talk tomorrow.” Completely worn out from the journey, Wang Meng lay down on the bed and quickly drifted off to sleep.

The next morning, sunlight filtered through the gaps in the wooden walls and fell across her face. She sat up slowly. Suddenly, she heard a faint rustling sound from the corner of the room. Startled, she turned to see a little girl quietly sweeping the floor with her head lowered. The girl looked up timidly. Pointing to herself, Wang Meng smiled. “I’m Wang Meng.” The little girl pointed to herself in return. “I’m Xiaomei,” she said softly.

“Where’s Old Chen?” Wang Meng asked, gesturing. Xiao Mei nodded and motioned for her to follow. Wang Meng stepped outside with the girl and saw Old Chen at the village entrance, talking with several people dressed in local attire. When Old Chen noticed her, he said something to the others, and they all turned to look at her. Wang Meng felt a little self‑conscious.

“Good morning, Dr. Wang Meng!” Old Chen said warmly. “Let me take you to see the clinic.”

“Yes, please. Thank you,” she replied quickly, her heart full of anticipation.

Old Chen led her along a winding path to the clinic. When they pushed open the door, she saw two simple plank beds laid directly on the floor, and a table cobbled together from wooden boards. The only medical equipment was an old‑fashioned stethoscope, a thermometer, and a nearly empty box of gauze. “But……Dr. Li Wei said the clinic was well‑stocked with supplies,” Wang Meng said, puzzled.

“Dr. Li Wei?” Old Chen repeated, looking confused. “Oh, right, right—Li Wei. The organization promised to send us supplies, but they never arrived. You know how it is—the mountain roads make deliveries slow, and things often get held up.”“It’s all right,” Wang Meng said. Until the new shipment came, she would have to rely on her own small medical kit to treat patients.

Xiao Mei soon became Wang Meng’s little companion. Every morning, she brought breakfast to the cabin. In the evenings, after Wang Meng returned from the clinic, Xiao Mei would help her tidy up. Xiao Mei taught Wang Meng the local dialect, and in return, Wang Meng began teaching Xiao Mei Mandarin.

The first week at the clinic went smoothly enough. Most of the villagers who came had only minor ailments. Wang Meng suspected that many weren’t really sick—they were just curious about the outsider. Still, she didn’t mind; she found them all rather endearing. Most of the villagers spoke only the local dialect, and when Old Chen was around, he could interpret. But more often than not, Wang Meng had to rely on gestures and facial expressions. The villagers were grateful for her help, and she felt a real sense of accomplishment.

One week in, an elderly woman named A Cai came to the clinic with a high fever and strange red rashes on her chest and back. Wang Meng prescribed medication, but her condition did not improve. That evening, Old Chen and another villager arrived at the clinic, but they stood at the door, reluctant to come closer, as if afraid of catching something. Wang Meng asked Old Chen if anyone in the village had ever shown similar symptoms. Old Chen shook his head no, but the other man’s face flickered with an odd expression. The next day, A Cai was gone. Xiao Mei said she had been taken away, but refused to say more.

In the second week, the village held a lively festival. The villagers sang and danced, their laughter echoing through the valleys. Old Chen sat beside Wang Meng and said gratefully, “Everyone in the village is so thankful that you’ve come all this way to treat us. We truly don’t know what we’d do without you.” Wang Meng felt a warm glow inside, happy to be part of this adventure.

Despite her contentment, she occasionally missed home. One morning, she discovered a faint mobile signal on the small hill behind the cabin. She quickly called her best friend Lin Lin and poured out all she had seen and heard in the mountain village, including the strange fever case.

After hanging up, Wang Meng went back to the cabin to rest for the afternoon. A knock came at the door. Old Chen entered hurriedly: “We’ve found loggers cutting trees in the forbidden zone. We need to contact the forestry department right away, but the satellite phone is broken. Could I borrow your mobile?” Wang Meng handed him her phone and charger. “Take your time, no rush,”she said. After Old Chen left, a young man came to the clinic with a fever, a rash, and a severe cough. Wang Meng stayed with him until late into the night, but he passed away. Even though she knew she had done all she could, grief and guilt overwhelmed her.

A few days later, she encountered her third strange fever case—A Qiang, a respected clan leader in the village and a close friend of Old Chen. Fortunately, his illness was not severe. Wang Meng treated him with medications and herbal remedies, and he eventually recovered. A Qiang’s family brought her many local specialties as thanks. Wang Meng finally felt that she had truly become part of this new community.

Still, Wang Meng remained frustrated by the villagers who had suffered from the fevers. She racked her brain for possible effective drugs and drew up a long list of medicines to give to Old Chen. Old Chen had not yet returned her phone, but she didn’t want to press him.

The next morning, on her way to the clinic, Wang Meng saw Old Chen standing in the center of the village, talking with those clan‑leader types. When she approached, the men abruptly fell silent. Wang Meng felt she might have intruded on something important. She handed the medicine list to Old Chen, who told her he would place the order as soon as possible. But then Wang Meng recalled that the earlier shipment he had promised had never arrived. That night, she tossed and turned in bed, unable to sleep. Her mind churned with questions: Could this be a disease never seen before? Sometimes she felt the villagers were hiding something from her. Did they know more about these fevers than they let on? Were they testing her medical skills? Or did they think that only by falling sick could they get medical care? But quickly, she felt ashamed of such thoughts. The villagers had been warm and friendly; there was no reason to believe they had known about the fevers before she came, and Dr. Li Wei had never mentioned anything about it in his email.

While lost in these anxious ruminations, Wang Meng suddenly realized that Xiao Mei had not come to the cabin that evening. She guessed the girl was busy helping her family prepare for the rainy season. She missed Xiao Mei’s company, but let it go.

The next morning at breakfast, Xiao Mei still did not appear. Wang Meng was busy at the clinic all day, and when she returned to the cabin in the evening, there was no sign of her. Worried and lonely, Wang Meng decided to go to Xiao Mei’s home.

She had never been inside Xiao Mei’s cabin, though she knew which one it was. She picked her way carefully across the clearing, avoiding the puddles left by the rain. She knocked softly on the door, but there was no answer. She didn’t want to intrude, and was about to turn away, but concern and curiosity made her pause. Finally, she pushed the door open.

What she saw made her freeze in shock. Xiao Mei lay on her bed, coughing uncontrollably, her body writhing in distress. A woman and a man were bent over her, trying to feed her a bowl of herbal concoction. Xiao Mei’s shirt had been removed, and by the dim glow of the fire, Wang Meng could clearly see the red rashes spreading across her chest. She drew a sharp breath, then looked up—and saw that the man was Old Chen, his eyes brimming with tears.

“Xiao Mei is your daughter?” Wang Meng asked, stunned. How could she not have known? Old Chen nodded, his face etched with despair.

“I’ll be right back,” Wang Meng said, and turned to dash to the clinic for the last of the fever medication. But as she spun around, she caught sight of her mobile phone lying on the table by the door. On instinct, she grabbed it and ran.

She did not head straight to the clinic. Instead, she took the oil lamp and hurried along the path leading down to the river. Gasping for breath, she reached the riverbank and sat down on a rock, staring blankly at the murky water. She longed for a boat to pass by, but the silence was absolute. At that moment, the full reality of her isolation struck her—she was in a place so remote, so cut off from the rest of the world.

How long had Xiao Mei been ill? Why had she kept it from her? Wang Meng suddenly remembered the hands that had prepared her breakfast, folded her clothes, and made her bed—and a chilling thought seized her: what if the disease was contagious? What if she herself had already been infected?

Clutching the phone in her hand, she tried to calm herself, but anxiety and fear spread like wildfire. A desperate urge to flee rose within her—to get away from this place and the unseen threat. Yet how could she abandon the villagers, especially the trusting little Xiao Mei, to such a fate? But if she stayed, facing this unknown illness, she had no idea whether she had the skill to cure it—or whether she would even survive.

Deep in agonizing conflict, Wang Meng was torn, unable to decide which path to take.

| **Table S1. Group Comparison of Intersubject Correlations(ISC)** | | | | | | |
| --- | --- | --- | --- | --- | --- | --- |
| Region | | | Peak (MNI) x, y, z | | ISC value | |
| **Parietal,Temporal, Occipital** | | | |  |  | |
| Occipital_Sup_R | | | 24, -68, 20 | | -0.01 | |
| Lingual_R | | | 16, -42, 0 | | -0.01 | |
| Cuneus_R | | | 20, -68, 20 | | -0.01 | |
| Calcarine_R | | | 4, -64, 18 | | -0.02 | |
| Occipital_Sup_L | | | -20, -80, 44 | | -0.02 | |
| Lingual_L | | | -12, -46, 0 | | -0.02 | |
| Calcarine_L | | | -12, -52, 6 | | -0.02 | |
| Temporal_Mid_R | | | 52, -28, -4 | | -0.1 | |
| Temporal_Sup_R | | | 64, -4, -4 | | -0.11 | |
| Temporal_Pole_Sup_R | | | 58, 6, -14 | | -0.1 | |
| Temporal_Mid_L | | | -54, -20, -10 | | -0.09 | |
| Temporal_Sup_L | | | 58, -12, -8 | | -0.11 | |
| Temporal_Inf_L | | | -42, 4, -34 | | -0.05 | |
| Temporal_Pole_Sup_L | | | -48, 8, -6 | | -0.07 | |
| Temporal_Pole_Mid_L | | | -36, 22, -34 | | -0.05 | |
| Heschl_R | | | 38, -22, 8 | | -0.01 | |
| Heschl_L | | | -34, -24, 8 | | -0.02 | |
| Parietal_Inf_R | | | 54, -36, 50 | | -0.02 | |
| Parietal_Inf_L | | | -24, -82, 44 | | -0.02 | |
| Parietal_Inf_L | | | -46, -42, 44 | | -0.02 | |
| Parietal_Sup_L | | | -20, -74, 44 | | -0.02 | |
| Precuneus_R | | | 12，-56，44 | | -0.03 | |
| Precuneus_L | | | -18, -48, 2 | | -0.02 | |
| SupraMarginal_R | | | 54, -44, 36 | | -0.03 | |
| SupraMarginal_L | | | -54, -46, 30 | | -0.04 | |
| Angular_R | | | 54, -50, 32 | | -0.06 | |
| Angular_L | | | -56, -58, 30 | | -0.02 | |
| Cingulate_Mid_L | | | -6, -38, 40 | | -0.02 | |
| Cingulate_Post_L | | | -4, -48, 22 | | -0.02 | |
| Fusiform_R | | | 42, -20, -26 | | -0.01 | |
| Fusiform_L | | | -22, -42, -14 | | -0.01 | |
| Cuneus_R | | | 24, -56, 24 | | -0.02 | |
| Cuneus_L | | | -6, -68, 26 | | -0.02 | |
| Occipital_Mid_R | | | 44, -78, 24 | | -0.02 | |
| Parietal_Sup_R | | | 14, -66, 54 | | -0.02 | |
| Insula_R | | | 44, 20, 0 | | -0.05 | |
| Insula_L | | | -46, 2, -4 | | -0.07 | |
| Hippocampus_R | | | 28, -30, -6 | | -0.01 | |
| Hippocampus_L | | | -20, -12, -14 | | -0.01 | |
| Precentral_R | | | 64, 10, 18 | | -0.02 | |
| Precentral_L | | | -58, 2, 26 | | -0.01 | |
| Postcentral_R | | | 62, 2, 16 | | -0.02 | |
| Postcentral_L | | | -58, -20, 30 | | -0.02 | |
| ParaHippocampal_R | | | 28, -28, -16 | | -0.02 | |
| ParaHippocampal_L | | | -16, 0, -24 | | -0.01 | |
| **Frontal** | | |  | |  | |
| Rolandic_Oper_R | | | 54, 4, 6 | | -0.02 | |
| Rolandic_Oper_L | | | -48, -22, 22 | | -0.02 | |
| Frontal_Sup_Medial_R | | | 8, 56, 22 | | -0.02 | |
| Frontal_Sup_Medial_L | | | -4, 64, 0 | | -0.02 | |
| Frontal_Mid_2_R | | | 36, 46, 2 | | -0.01 | |
| Frontal_Mid_2_L | | | -40, 32, 32 | | -0.01 | |
| Frontal_Med_Orb_R | | | 4, 34, -12 | | -0.02 | |
| Frontal_Med_Orb_L | | | -6, 42, -12 | | -0.02 | |
| Frontal_Inf_Tri_R | | | 52, 28, 0 | | -0.05 | |
| Frontal_Inf_Tri_L | | | -44, 24, 2 | | -0.02 | |
| Frontal_Inf_Oper_R | | | 50, 18, 6 | | -0.05 | |
| Frontal_Inf_Oper_L | | | -40, 20, 32 | | -0.02 | |
| Frontal_Inf_Orb_2_R | | | 50, 38, -12 | | -0.04 | |
| Frontal_Inf_Orb_2_L | | | -30, 28, -8 | | -0.02 | |
| OFCpost_R | | | 40, 22, -14 | | -0.03 | |
| OFCpost_L | | | -36, 22, -14 | | -0.02 | |
| OFClat_R | | | 50, 38, -14 | | -0.04 | |
| OFClat_L | | | -48, 24, -14 | | -0.02 | |
| OFCmed_L | | | -18, 28, -16 | | -0.01 | |
| Rectus_R | | | 4, 30, -16 | | -0.02 | |
| Rectus_L | | | 2, 30, -16 | | -0.02 | |
| Olfactory_R | | | 4, 20, -8 | | -0.02 | |
| Olfactory_L | | | -6, 24, -10 | | -0.01 | |
| ACC_pre_R | | | 6, 38, 18 | | -0.02 | |
| ACC_pre_L | | | -6, 50, 0 | | -0.02 | |
| ACC_sup_R | | | 6, 28, 24 | | -0.02 | |
| ACC_sub_R | | | 4, 24, -8 | | -0.02 | |
| ACC_sub_L | | | -4, 28, -10 | | -0.01 | |
| Cingulate_Mid_R | | | 8, 36, 30 | | -0.02 | |
| Cingulate_Post_R | | | 4, -50, 24 | | -0.02 | |
| Frontal_Sup_2_R | | | 24, 18, 44 | | -0.02 | |
| Frontal_Sup_2_L | | | -20, 28, 40 | | -0.01 | |
| ACC_sup_L | | | -4, 34, 20 | | -0.02 | |
| Supp_Motor_Area_L | | | -6, 4, 46 | | -0.02 | |
| **Subcortical/Cerebellum** | | |  | |  | |
| Cerebellum_3_R | | | 12, -40, -8 | | -0.01 | |
| Cerebellum_4_5_R | | | 12, -42, -8 | | -0.01 | |
| Cerebellum_8_L | | | -32, -64,-52 | | -0.01 | |
| Cerebellum_7b_L | | | -38, -64, -52 | | -0.01 | |
| Putamen_R | | | 30, 16, 6 | | -0.02 | |
| Amygdala_L | | | -24, -6, -14 | | -0.01 | |
| Cerebellum_Crus2_L | | | -40,-60,-50 | | -0.01 | |
| Cerebellum_8_R | | | 16, -66, -42 | | -0.01 | |
| Cerebellum_4_5_L | | | -14,-46,-12 | | -0.01 | |
| **Table S2 Group Comparison of IS-RSA (Totalscore)** | | | | | | |
| Region | Peak (MNI) x, y, z | | | | | IS-RSA value |
| **Parietal ,Temporal,Occipital** | |  | | | |  |
| Temporal_Sup_R | | 44, -36, 4 | | | | 0.37 |
| Temporal_Inf_R | | 44, -50, -16 | | | | 0.14 |
| Temporal_Mid_R | | 52, -30, -8 | | | | 0.37 |
| Insula_R | | 40, -18, 8 | | | | 0.25 |
| Insula_L | | -36, 16, -6 | | | | 0.15 |
| Heschl_R | | 38, -22, 8 | | | | 0.25 |
| Angular_R | | 58, -62, 24 | | | | 0.2 |
| Occipital_Mid_R | | 54, -66, 24 | | | | 0.2 |
| Occipital_Sup_R | | 28, -72, 46 | | | | 0.13 |
| Occipital_Inf_R | | 50, -76, -2 | | | | 0.17 |
| Fusiform_R | | 44, -40, -16 | | | | 0.14 |
| Parietal_Inf_R | | 56, -48, 46 | | | | 0.15 |
| SupraMarginal_R | | 62, -50, 42 | | | | 0.15 |
| ParaHippocampal_R | | 20, 4, -24 | | | | 0.15 |
| Temporal_Pole_Sup_R | | 40, 20, -24 | | | | 0.17 |
| Temporal_Pole_Mid_R | | 50, 8, -24 | | | | 0.19 |
| Temporal_Pole_Sup_L | | -50, 18, -18 | | | | 0.21 |
| Temporal_Pole_Mid_L | | -46, 18, -26 | | | | 0.21 |
| Temporal_Mid_L | | -16, 2, -18 | | | | 0.16 |
| Temporal_Inf_L | | -52, -10, -26 | | | | 0.16 |
| Temporal_Sup_L | | -56, -44, 18 | | | | 0.22 |
| ParaHippocampal_L | | -18, -36, -12 | | | | 0.16 |
| Fusiform_L | | -18, -32, -18 | | | | 0.16 |
| Lingual_L | | -18, -46, -6 | | | | 0.19 |
| SupraMarginal_L | | -56, -44, 30 | | | | 0.23 |
| Angular_L | | -54, -58, 30 | | | | 0.15 |
| Parietal_Inf_L | | -58, -52, 42 | | | | 0.18 |
| Fusiform_R | | 46,-58,-20 | | | | 0.14 |
| Lingual_R | | 26,-56,-4 | | | | 0.11 |
| Parietal_Sup_R | | 52,-54,52 | | | | 0.15 |
| Heschl_L | | -36,-26,8 | | | | 0.16 |
| ParaHippocampal_L | | -26, -8, -26 | | | | 0.13 |
| Precentral_L | | -42, 12, 32 | | | | 0.13 |
| Postcentral_R | | 50, -18, 36 | | | | 0.1 |
| **Frontal** | |  | | | |  |
| Frontal_Sup_2_R | | 22, 64, 2 | | | | 0.14 |
| Frontal_Mid_2_R | | 32, 54, -2 | | | | 0.14 |
| Frontal_Inf_Tri_R | | 44, 26, 20 | | | | 0.13 |
| Frontal_Inf_Orb_2_R | | 40, 48, -2 | | | | 0.12 |
| OFClat | | 48, 44, -14 | | | | 0.12 |
| OFCant | | 40, 48, -14 | | | | 0.12 |
| Frontal_Med_Orb_R | | 14, 62, -8 | | | | 0.14 |
| Frontal_Inf_Tri_L | | -42, 14, 24 | | | | 0.13 |
| Frontal_Inf_Oper_L | | -36, 18, 32 | | | | 0.13 |
| Frontal_Inf_Orb_2_L | | -54, 34, -6 | | | | 0.13 |
| Frontal_Mid_2_L | | -44, 22, 32 | | | | 0.14 |
| OFClat_L | | -44, 36, -18 | | | | 0.13 |
| OFCpost_L | | -40, 34, -20 | | | | 0.13 |
| OFClat_R | | 42,44,-20 | | | | 0.12 |
| OFCant_R | | 42,46,-14 | | | | 0.12 |
| Rolandic_Oper_L | | -34,-26,14 | | | | 0.16 |
| Rolandic_Oper_R | | 50, -8, 12 | | | | 0.2 |
| Cingulate_Mid_R | | 4, -24, 38 | | | | 0.16 |
| **Subcortical/Cerebellum** | |  | | | |  |
| Cerebellum_9_R | | 4, -58, -52 | | | | 0.15 |
| Cerebellum_7b_R | | 40, -70, -52 | | | | 0.13 |
| Cerebellum_8_R | | 8, -68, -32 | | | | 0.13 |
| Cerebellum_Crus2_R | | 6,-68,-30 | | | | 0.13 |
| Cerebellum_4_5_R | | 24, -30, -24 | | | | 0.13 |
| Cerebellum_Crus1_R | | 48, -68, -20 | | | | 0.14 |
| Amygdala_R | | 24, 4, -20 | | | | 0.16 |
| Putamen_L | | -30, -8, 4 | | | | 0.12 |
| Hippocampus_L | | -24,-10,-26 | | | | 0.13 |
| Cerebellum_4_5_L | | -18, -36, -18 | | | | 0.16 |
| Cerebellum_Crus1_L | | -34, -80, -30 | | | | 0.15 |
| Cerebellum_Crus2_L | | -20, -90, -30 | | | | 0.15 |
| Vermis_9 | | 0, -54, -30 | | | | 0.12 |
| Cerebellum_9_L | | -12, -58, -54 | | | | 0.14 |
| Cerebellum_8_L | | -22, -52, -54 | | | | 0.14 |
| Vermis_8 | | 2,-62,-36 | | | | 0.14 |

| **Table S3 Group Comparison of IS-RSA (Persecution)** | | | |
| --- | --- | --- | --- |
| Region | | Peak (MNI) x, y, z | IS-RSA value |
| **Parietal ,Temporal,Occipital** |  | |  |
| Angular_R | 42, -54, 22 | | 0.2 |
| Temporal_Sup_R | 46, -30, -2 | | 0.34 |
| Temporal_Inf_R | 58, -52, -6 | | 0.21 |
| Temporal_Mid_R | 52, -36, -2 | | 0.34 |
| Occipital_Mid_R | 53, -78, -2 | | 0.17 |
| Occipital_Inf_R | 48, -78, -2 | | 0.17 |
| Fusiform_R | 50, -66, -18 | | 0.15 |
| ParaHippocampal_R | 22, -24, -18 | | 0.15 |
| Heschl_R | 36, -26, 6 | | 0.21 |
| Insula_R | 38, -22, 6 | | 0.21 |
| Parietal_Inf_R | 46, -56, 46 | | 0.18 |
| SupraMarginal_R | 50, -38, 38 | | 0.14 |
| Cuneus_R | 12, -98, 6 | | -0.1 |
| Occipital_Sup_R | 18, -98, 6 | | -0.1 |
| Calcarine_R | 10, -94, 0 | | -0.1 |
| Lingual_R | 6, -88, -12 | | -0.1 |
| Temporal_Pole_Mid_R | 34, 12, -34 | | 0.12 |
| Temporal_Pole_Sup_R | 40, 14, -28 | | 0.12 |
| Lingual_L | -12, -42, -8 | | 0.16 |
| Temporal_Pole_Mid_L | -40, 14, -38 | | 0.18 |
| Temporal_Pole_Sup_L | -36, 18, -28 | | 0.18 |
| Temporal_Inf_L | -48, 8, -38 | | 0.18 |
| Temporal_Sup_L | -60, -46, 18 | | 0.21 |
| Temporal_Mid_L | -62, -2, -22 | | 0.18 |
| SupraMarginal_L | -52, -46, 26 | | 0.21 |
| Angular_L | -56, -58, 26 | | 0.16 |
| Parietal_Inf_L | -46, -48, 60 | | 0.2 |
| Fusiform_L | -16, -46, -12 | | 0.16 |
| Insula_L | -34, 14, -4 | | 0.15 |
| ParaHippocampal_L | -24,-42,-8 | | 0.14 |
| **Frontal** |  | |  |
| Frontal_Med_Orb_R | 14, 62, -8 | | 0.16 |
| Frontal_Sup_2_R | 20, 62, -8 | | 0.16 |
| Frontal_Mid_2_R | 32, 52, -2 | | 0.16 |
| Frontal_Inf_Tri_R | 48,26,20 | | 0.15 |
| Frontal_Inf_Orb_2_R | 46, 44, -4 | | 0.15 |
| Frontal_Inf_Oper_R | 58, 22, 28 | | 0.15 |
| OFClat_R | 44, 42, -14 | | 0.15 |
| OFCant_R | 42, 48, -14 | | 0.15 |
| Frontal_Sup_Medial_R | 6, 32, 48 | | 0.12 |
| Supp_Motor_Area_R | 6, 22, 48 | | 0.12 |
| Frontal_Inf_Orb_2_L | -40, 30, -4 | | 0.15 |
| Frontal_Inf_Oper_L | -46, 14, 22 | | 0.15 |
| Frontal_Mid_2_L | -28, 20, 60 | | 0.13 |
| Frontal_Sup_2_L | -24, 22, 60 | | 0.13 |
| Frontal_Inf_Tri_L | -46, 18, 22 | | 0.15 |
| OFCant_L | -30, 36, -14 | | 0.11 |
| OFCpost_L | -34, 32, -14 | | 0.11 |
| OFClat_L | -42, 32, -16 | | 0.15 |
| Rectus_R | 4,24,-24 | | -0.12 |
| Olfactory_R | 4,12,-14 | | -0.12 |
| Cingulate_Mid_L | -12,-26,42 | | 0.16 |
| Frontal_Sup_Medial_L | -6,48,46 | | 0.12 |
| Rolandic_Oper_R | 46, -14, 12 | | 0.2 |
| Cingulate_Mid_R | 2, -30, 46 | | 0.16 |
| **Subcortical/Cerebellum** |  | |  |
| Hippocampus_L | -16,-28,-12 | | 0.14 |
| Cerebellum_Crus1_R | 48, -68, -20 | | 0.15 |
| Cerebellum_8_R | 14, -70, -44 | | 0.13 |
| Cerebellum_7b_R | 10, -74, -44 | | 0.13 |
| Cerebellum_9_R | 2, -60, -44 | | 0.14 |
| Cerebellum_4_5_R | 24, -26, -26 | | 0.15 |
| Vermis_8 | 4, -74, -40 | | 0.13 |
| Vermis_9 | 2, -62, -44 | | 0.14 |
| Putamen_L | -24, -8, 12 | | 0.12 |
| Pallidum_L | -26, -10, -4 | | 0.12 |
| Cerebellum_4_5_L | -14, -46, -12 | | 0.16 |
| Cerebellum_Crus1_L | -12, -84, -24 | | 0.15 |
| Cerebellum_Crus2_L | -12, -80, -30 | | 0.15 |

| **Table S4 Group Comparison of IS-RSA (Reference)** | | | |
| --- | --- | --- | --- |
| Region | | Peak (MNI) x, y, z | IS-RSA value |
| **Parietal ,Temporal,Occipital** |  | |  |
| Parietal_Inf_R | 40, -50, 40 | | 0.1 |
| Angular_R | 46, -62, 26 | | 0.19 |
| Occipital_Mid_R | 46, -64, 24 | | 0.19 |
| Occipital_Inf_R | 34, -86, -6 | | 0.16 |
| Temporal_Inf_R | 56, 6, -36 | | 0.22 |
| Temporal_Mid_R | 58, -58, 18 | | 0.26 |
| Temporal_Sup_R | 50, -40, 22 | | 0.33 |
| SupraMarginal_R | 68, -26, 18 | | 0.35 |
| Insula_R | 36, -14, 12 | | 0.24 |
| Fusiform_R | 32, -40, -12 | | 0.13 |
| Lingual_R | 20, -60, -4 | | 0.13 |
| Temporal_Pole_Sup_R | 34, 8, -24 | | 0.17 |
| Temporal_Pole_Mid_R | 54, 14, -26 | | 0.22 |
| Heschl_R | 40, -22, 8 | | 0.24 |
| Temporal_Pole_Mid_L | -46, 8, -32 | | 0.18 |
| Temporal_Pole_Sup_L | -40, 22, -32 | | 0.18 |
| Temporal_Inf_L | -42, 4, -36 | | 0.18 |
| Temporal_Sup_L | -60, -34, 16 | | 0.24 |
| Temporal_Mid_L | -52, -34, 8 | | 0.24 |
| Insula_L | -40, 16, -8 | | 0.15 |
| ParaHippocampal_L | -26, -42, -6 | | 0.18 |
| Lingual_L | -22, -44, -6 | | 0.18 |
| Postcentral_R | 50,-16,36 | | 0.09 |
| Heschl_L | 34,-26,6 | | 0.17 |
| Parietal_Inf_L | -42, -56, 52 | | 0.11 |
| Occipital_Sup_L | -22,-88,22 | | 0.1 |
| Parietal_Inf_L | -46,-54,42 | | 0.14 |
| SupraMarginal_L | -60, -40, 24 | | 0.18 |
| Angular_L | -56, -56, 24 | | 0.11 |
| Fusiform_L | -18, -46, -12 | | 0.18 |
| **Frontal** |  | |  |
| Frontal_Sup_2_R | 20, 64, -2 | | 0.1 |
| Frontal_Mid_2_R | 30, 52, -2 | | 0.1 |
| Frontal_Med_Orb_R | 14,62,-4 | | 0.1 |
| Frontal_Inf_Oper_R | 52,12,8 | | 0.11 |
| OFClat_L | -46,32,-16 | | 0.12 |
| Rolandic_Oper_L | -40,-32,16 | | 0.17 |
| Rolandic_Oper_R | 48, -6, 12 | | 0.15 |
| OFCpost_R | 34, 16, -24 | | 0.17 |
| Frontal_Inf_Orb_2_L | -42,24,-4 | | 0.12 |
| **Subcortical/Cerebellum** |  | |  |
| Cerebellum_9_R | 8, -52, -40 | | 0.18 |
| Cerebellum_8_R | 8, -64, -42 | | 0.13 |
| Vermis_9 | 2, -54, -30 | | 0.13 |
| Vermis_8 | 4, -66, -42 | | 0.13 |
| Hippocampus_L | -14, -8, -20 | | 0.16 |
| Thal_PuM_L | -10, -32, 2 | | 0.16 |
| Thal_MDm_L | -4, -20, 2 | | 0.15 |
| Thal_IL_L | -12, -18, 2 | | 0.15 |
| Thal_VPL_L | -16, -18, 2 | | 0.15 |
| Thal_PuI_L | -16, -30, 2 | | 0.16 |
| Thal_PuA_L | -14, -26, 2 | | 0.16 |
| Thal_MGN_L | -16, -28, 0 | | 0.16 |
| Thal_VL_L | -16, -16, 2 | | 0.15 |
| Cerebellum_Crus1_L | -14, -84, -22 | | 0.15 |
| Cerebellum_6_L | -38, -68, -22 | | 0.15 |
| Cerebellum_Crus2_L | -14, -82, -30 | | 0.15 |
| Cerebellum_8_L | -14, -58, -58 | | 0.15 |
| Cerebellum_9_L | -10, -46, -58 | | 0.15 |
